# There Goes the Neighborhood? The Public Safety Enhancing Effects of a Mobile Harm Reduction Intervention

**DOI:** 10.1101/2023.05.30.23290739

**Authors:** Alex L. Fixler, Leah A. Jacobs, Daniel B. Jones, Aaron Arnold, Emily E. Underwood

## Abstract

**Aims:** To estimate the impact of mobile clinics providing medication for opioid use disorders on neighborhood arrest rates.

**Design:** A quasi-experimental difference-in-differences estimation.

**Setting:** Pittsburgh, Pennsylvania.

**Participants:** Census blocks in the 1-mile circumferences surrounding 4 mobile medication clinics.

**Intervention and comparators:** The intervention is comprised of mobile clinics providing buprenorphine to community members with opioid use disorders. A treatment group of eighty-four census block groups in the immediate areas surrounding clinics during the time period after their establishment were compared to a control group of city census blocks not within 1 mile of a clinic plus treated census block groups in the two years prior to clinic establishment.

**Measurements:** Outcome variables include drug, non-drug, and total arrests, measured per 100 in population.

**Findings:** Compared to block groups further than 1 mile from an MMC, we found that total arrests fell by 34.13% (*b* = -0.358, 95% CI = -0.557, -0.158), drug arrests fell by 33.85% (*b* = -0.087, 95% CI = -0.151, -0.023), and non-drug related arrests fell by 22.29% (*b* = -0.179, 95% CI = -0.302, -0.057). Drug arrests declined significantly on days when the MMCs were not present (b = -0.015, 95% CI = -0.025, -0.006), but did not change significantly on the days when the intervention was active and on site (*b* = -0.002, 95% CI = -0.016, -0.013). Total arrests declined significantly on days when MMCs were and were not present (*b* = -0.045, 95% CI = - 0.078, -0.012; and *b* = -0.052, CI = -0.082, -0.023, respectively).

**Conclusions:** Mobile clinics providing medication for opioid use disorders significantly reduced neighborhood arrest rates. Expansion of mobile services could promote health equity and public safety.

## INTRODUCTION

Medications for Opioid Use Disorder (MOUD), including methadone and buprenorphine, are associated with sizable reductions in drug poisonings and all-cause mortality (for a review, see Santo et al., 2021 [1]). Given these benefits and its place as a relatively low-barrier form of MOUD, buprenorphine has become a recommended treatment for opioid use disorder (OUD). Yet, utilization of buprenorphine is rare among people with OUD (2), and access is unequally distributed by race, income, and geography (3,4).

Several structural and social factors impede access to and use of buprenorphine. Though buprenorphine can be prescribed via virtual appointments and can be taken without daily direct observation (unlike methadone), other regulations have limited the availability of waivered clinicians and require frequent visits with providers for refills (5). Some of these structural barriers to care have been reduced, and have led to increased buprenorphine use (6). However, social impediments remain. “Not in my backyard” (i.e., NIMBY) resistance to substance use service sites is one such impediment (7,8). In low-income, African American and other communities of color, NIMBY sentiments are compounded by legacies of governmental and industrial placement of unsalutary and undesirable infrastructure and institutions (e.g., highways, waste management facilities, and correctional facilities). In other words, unique challenges to expanding access to treatment exist in communities that may face the greatest need.

Chief among NIMBY concerns is that substance use services will increase crime rates where they are placed. Theoretically, by attracting people disproportionately involved in crime (people who use drugs illegally), services could increase criminal activity in surrounding areas. It is also possible that the presence of substance use services will decrease crime-- at an individual level, by reducing illicit drug use and drug-related arrests, or at an ecological level, whereby providers may act as sources of social control or surveillance thought to discourage crime more generally (9,10). Empirically, few studies speak directly to these arguments. Estimating effects on county crime rates, Bondurant and colleagues found substance use service presence predicted reductions in serious violent crimes, burglaries, and motor vehicle thefts (11). Assessing more localized effects, Boyd and colleagues found methadone clinics were not associated with overall crime rates in their immediate surroundings (12). These studies suggest NIMBY concerns may be unwarranted. They do not, however, speak to the impact of recent innovations in service delivery that are intended to increase access but nonetheless face opposition—i.e., low-barrier MOUD and non-brick and mortar interventions, such as buprenorphine mobile services.

This study addresses concerns regarding the placement of substance use services and neighborhood crime rates. It does so by estimating the causal effect of a low-barrier, mobile MOUD intervention on general and specific crime. Results have important implications for theorizing the relationship between substance use services and crime, and efforts to expand access to MOUD in neighborhoods that face significant need.

## METHODS

### Intervention

Mobile MOUD clinics (MMC), initiated by harm reduction health service provider Prevention Point Pittsburgh (PPP), provide buprenorphine access alongside PPP’s syringe service program. The MMCs operate for 3 hours weekly per site. They are located in Pittsburgh neighborhoods in the north, south, east, and center of the city, and initiated service on different dates. Two were established in the fourth quarter of 2020, and two more in the first quarter of 2021. PPP determined intervention locations strategically to expand access to services based on evidence of need in the community.

The intervention aims to expand the availability of buprenorphine and create a community-based alternative to existing MOUD structures. By co-locating the MMCs with an existing syringe service program, the MMCs work to build a unique continuum of care for people seeking to reduce the harms of substance use. Service delivery is supportive and non-punitive, appointments are not necessary to receive care, and all out-of-pocket pharmacy costs are paid by the program, which also provides transportation support. The program is staffed by a social worker and a team of contracted medical providers from a regional health system. To receive services, clients first register with the social worker and identify treatment needs and goals. Next, they meet with a medical provider to discuss substance use history and eligibility for starting or continuing buprenorphine. If appropriate, the provider submits a prescription to the patient’s pharmacy of choice for same-day pickup. Services are delivered from a large cargo van that has been converted to include the core features of a clinic room. See Appendix A in our supplementary materials for more information about the intervention.

### Design

The study uses a quasi-experimental design to identify the causal impact of mobile MOUD clinics on local arrest rates. We utilize a staggered difference-in-differences estimation approach to infer this effect by comparing arrest trends in areas with and without MMCs prior to and after their establishment. This study was not pre-registered, and results should be considered exploratory.

### Data

To prepare the data for analysis, we identified the geographic coordinates of the MMCs, and used the *geodist* package in the statistical software Stata (13,14) to assign all census block groups within Pittsburgh city limits to the nearest MMC and also to identify all census block groups within one mile (*n* = 84) circumference of any MMC. Census blocks within these surrounding areas constitute “treated” units. Our control group included all blocks beyond one mile from an MMC (n = 205).

Our primary outcome variables are (per 100 in population) counts of arrests, both total and within specific offense categories, aggregated to the census block group-by-quarter level. Specifically, the raw data are drawn from the Pittsburgh Police Department’s public records of arrests (15). The raw data are incident-level administrative records that include the date, time, precise location, and alleged offense associated with each arrest, for the universe of arrests in the city. We used the location to attach incidents to census block groups to coincide with the “treated” and “control” areas described above and used the date of the incident to temporally group incidents into year-quarters. We then aggregated incidents (overall and by offense category) to the block group-by-quarter level. We also divided arrest counts by each 100 in the block group’s population to normalize across differently sized block groups.

Frequently, multiple offense codes were associated with individual arrests. We organized offense types into the following broad categories of interest: drug (e.g. possession or use of controlled substances or paraphernalia), property (e.g. theft or robbery), violent (i.e. the perpetration or threat of bodily harm), weapon (e.g. possessing an unlicensed firearm or use of a weapon alongside another offense), public order (e.g. disorderly conduct or loitering), and vehicular (e.g. unlicensed or impaired driving). Although the WPRDC uses a single umbrella code to categorize most drug-related charges, cannabis charges are coded separately. Since MOUD does not address cannabis misuse, we did not include cannabis charges in drug arrests (they are represented only when co-occurring with other drug-related charges). Since records did not otherwise differentiate between types of drugs involved in any arrest, all other drug charges were included in this category. Individual arrests commonly reflect multiple offense codes; in aggregating data to block group-by-quarter level, an individual arrest may contribute to counts of multiple categories. For example, an arrest with both a property and drug offense would contribute one additional arrest to: total arrests, property arrests, and drug arrests. Thus, the sum of arrests across the specific categories exceeds our count of total arrests (“All” in Table 1).

**Table 1.** Summary statistics: Averages of arrests, pre- and post-arrival of nearby MMC

|  | (1)<br>All | (2)<br>Pre-MMC<br>< 1 mi. | (3)<br>Post-MMC<br>< 1 mi. | (4)<br>Pre- MMC<br>> 1 mi. | (5)<br>Post-MMC<br>> 1 mi. |
| --- | --- | --- | --- | --- | --- |
| Overall | 0.98 | 1.51 | 0.94 | 0.90 | 0.86 |
| Drug | 0.42 | 0.60 | 0.25 | 0.49 | 0.30 |
| Property | 0.41 | 0.23 | 0.17 | 0.52 | 0.42 |
| Violent | 0.60 | 0.48 | 0.36 | 0.69 | 0.62 |
| Weapon | 0.11 | 0.09 | 0.11 | 0.09 | 0.16 |
| Pub. Order | 0.43 | 0.32 | 0.18 | 0.51 | 0.46 |
| Vehicle | 0.36 | 0.23 | 0.15 | 0.45 | 0.36 |
| Observations | 6669 | 902 | 656 | 2959 | 2152 |

*Note.* Incidents are aggregated to a block-group-by-quarter level and reported on a per 100 in population basis.

### Analysis

We used a difference-in-differences model to compare arrest trends in areas near MMCs to other areas in the city during the same time period, and account for the progressive introduction of the treatment. This is accomplished by including as dependent variables the interactions between indicator variables for being within one mile of an MMC (=1 if so, =0 if not) and an indicator variable “post” equal to 1 in the quarters including and following the arrival of a nearby MMC. We therefore captured potential “treatment” effects within a one-mile range of each, relative to a control set of block groups greater than one mile from the nearest MMC. Lacking an ex ante prediction as to the exact geographic extent of the impact of MMC’s, we chose the range based on initial analyses in which we tested for impacts at other distances, e.g. 0-0.5 miles and 1-1.5 miles. Based on those outcomes, we were able to narrow the range of the study to impacted locations with less concern for “bleed” from service users based outside of treatment areas (see Appendix B in supplementary materials). Consultation with PPP providers supported our perception that most effects would take place within the mile surrounding an MMC, as they confirmed that many clients traveled to clinics on foot, and that most participants lived in the immediate neighboring area. We also included an additional model to measure outcomes on different days of the week, based on the presence or absence of an MMC (controlling for potentially different arrest patterns on weekends).

We included block group fixed effects to account for time invariant characteristics of block groups and year-quarter fixed effects to control for factors impacting all block groups in the city across time -- COVID being a notable example of a factor captured by the year-quarter fixed effects. The former was important in capturing the fact that MMCs are not randomly allocated to neighborhoods and therefore there are some systematic differences in neighborhoods that receive MMCs versus those that do not. The block group fixed effects accounted for these differences.

Given the count nature of our data and the associated positive skew, we estimated all specifications as Poisson models. In most tables, we report the average marginal effects resulting from these estimates for ease of interpretation. Standard errors are clustered at block group-level.

Finally, critical to a difference-in-differences design is that trends across the treatment and control groups evolved similarly prior to the introduction of “treatment” (i.e., arrival of MMCs). To test that assumption, we conducted event studies to examine arrest rates in the intervention and comparison groups before and after MMC arrival, and identified any emerging trends that might have preceded them. The event study models are similar to the main difference-in-differences model, but instead of interacting the spatial proximity variable with a simple “Post” indicator, we interact with a full set of indicators capturing how many periods pre-or post-MMC arrival the relevant quarter is from. In doing so, we omit the two quarters immediately before van arrival to serve as the reference category (as the “pre” period does in the difference-in-differences.)

### Secondary analyses

We ran additional tests to probe the robustness of our main results. These tests included: (1) redefining the proximity to an MMC that defines “treatment” (0.5 miles, 1.5 miles); (2) restricting the sample to block groups within 1.5 miles to achieve a more comparable control group (assuming nearby block groups are more similar to those in close proximity to an MMC); (3) estimating a propensity score based on block group demographics that predicts the likelihood of being near an MMC and reweighting regressions using inverse probability weights; and (4) estimating our main specification as Ordinary Least Squares (OLS) rather than Poisson. These results are available in full in Appendix C.

## RESULTS

### Summary statistics

The means in Table 1 preview our main result: there is a large decrease in arrests post-MMC in block groups within 1 mile of an MMC, but not in block groups that are more distant. Arrest patterns in areas within one mile of MMCs varied from those outside a mile. Prior to the establishment of the MMCs, the average city arrest rate inside one mile of their eventual locations was 60 percent higher than those inside the immediate surrounding mile of each van. The areas inside this designated treatment region also had higher drug arrest rates, and lower property, violent, public order, and vehicle crime than those outside of it. After their establishment, the areas within one mile of the vans also had lower drug and weapons arrest rates than outside the treatment area, while total arrest rates were still slightly higher. See Table 1 for a detailed summary of arrest rates per 100 capita at a block-group-by-quarter level prior to and after the establishment of the MMCs, and Figure 1 for a scatterplot of drug arrests by proximity to MMCs prior to and after their arrivals.

**FIGURE 1.**
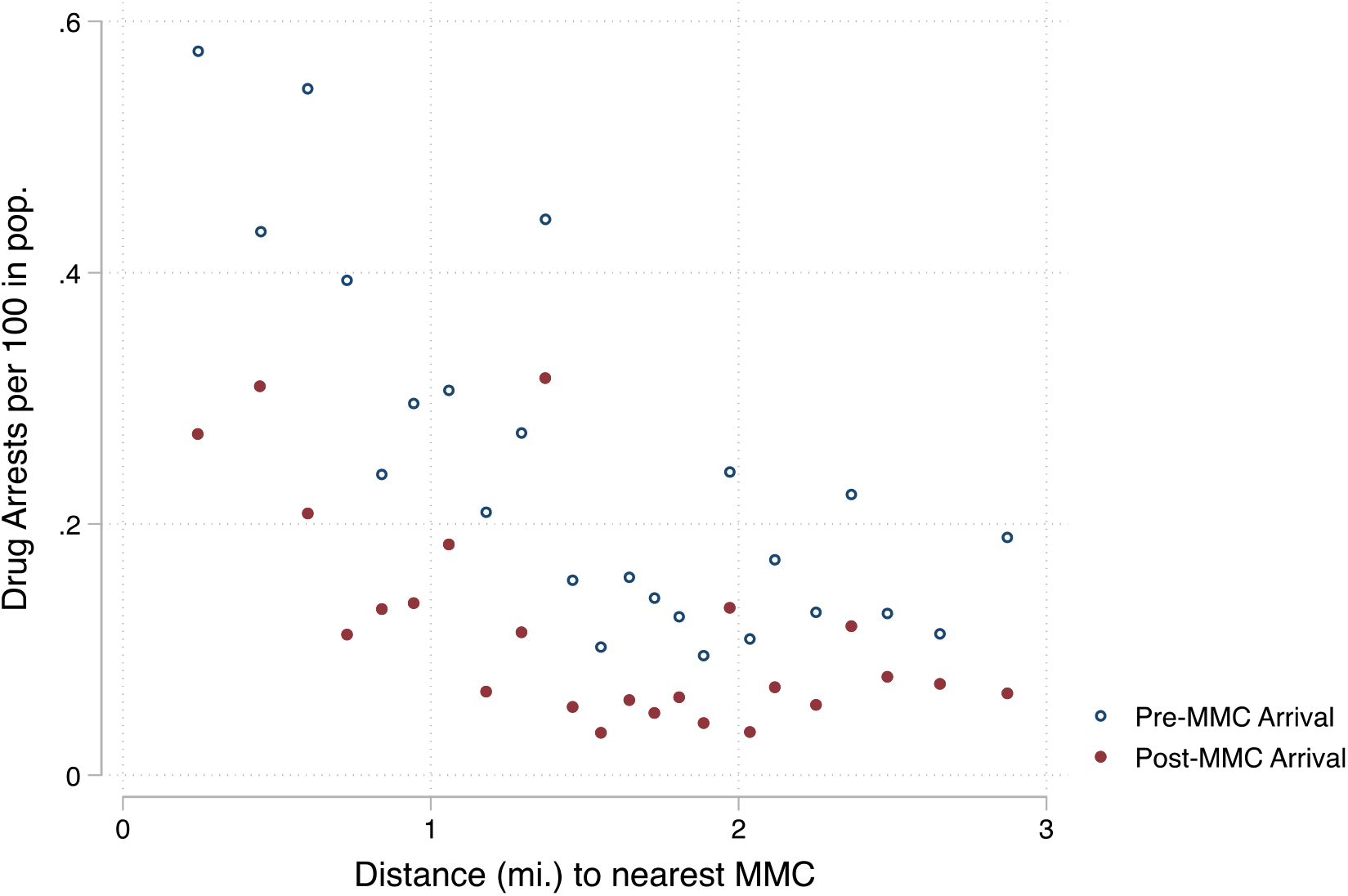
Scatterplot charting binned averages of drug arrests per 100 capita at different distances from MMC

### Primary regression outcomes

Given higher overall and drug arrest rates in the areas closer to the MMCs, our main specification measured the change in these categories in the mile surrounding MMCs. Additional specifications estimate effects in other arrest categories. All models control for block-by-quarter fixed effects. All outcomes are estimated as Poisson regressions; we report average marginal effects.

Compared to block groups further than 1 mile from an MMC, we found that total arrests fell by 0.358 per 100 in population (95% CI = -0.557, -0.158), drug arrests by 0.087 (95% CI = -0.151, - 0.023), and non-drug related arrests by 0.179 (95% CI = -0.302, -0.057) (see Table 2). The reduction in drug arrests was smallest in absolute terms, but the baseline average of that outcome is also the lowest (reported in the second to last row of Table 2); both Drug Arrests and Total Arrests declined by roughly 34% relative to their baseline average.

**Table 2.** Outcomes: Total arrests, drug arrests, and non-drug arrests per 100 in pop.

| VARIABLES | (1)<br>All | (2)<br>Drug | (3)<br>Not Drug |
| --- | --- | --- | --- |
| Post X 0-1 mi. | -0.358***<br>(-0.557 - -0.158) | -0.087***<br>(-0.151 - -0.023) | -0.179***<br>(-0.302 - -0.057) |
| Observations | 6,612 | 6,384 | 6,593 |
| Outcome avg. | 1.049 | 0.257 | 0.803 |
| Estimate p-value | 0.000 | 0.008 | 0.004 |

95% confidence intervals in parentheses below estimates. All columns estimated as Poisson regressions.
Table reports average marginal effects. Standard errors clustered at block group-level. “Estimate p-value” in bottom row is exact p-value associated with the main coefficient estimate in column.
\*\*\* $p < 0.01$ , \*\* $p < 0.05$ , \* $p < 0.1$

Subsequent analyses of non-drug arrest categories (property, violent, weapon, public order, and vehicle) indicated variation in effects – violent, public order and vehicle charges fell significantly (*b* = -0.120, 95% CI = -0.251, 0.011; *b* = -0.155, 95% CI = -0.255, -0.055; and *b* = -0.126, 95% CI = -0.216, -0.037, respectively), while property and weapons offenses dropped slightly but not statistically significantly. See Table 3 for more detail on arrest type breakdowns. Finally, we found that drug arrests declined significantly on days when the MMCs were not present (b = - 0.015, 95% CI = -0.025, -0.006), but did not appear to change significantly on the days when the intervention was active and on site. Total arrests declined significantly on days when MMCs both were and were not present (*b* = -0.045, 95% CI = -0.078, -0.012; and *b* = -0.052, CI = - 0.082, -0.023, respectively), as did non-drug arrests (*b* = -0.032, CI = -0.061, -0.002 when present and *b* = -0.025, CI = -0.043, -0.006 when not). Table 4 provides a more detailed breakdown of arrest outcomes based on the presence or absence of MMCs.

**Table 3.** Outcomes: Other arrest categories per 100 in pop.

| VARIABLES | (1)<br>Property | (2)<br>Violent | (3)<br>Weapon | (4)<br>Pub. Order | (5)<br>Vehicle |
| --- | --- | --- | --- | --- | --- |
| Post X 0-1 mi. | -0.027<br>(-0.078 - 0.025) | -0.120*<br>(-0.251 - 0.011) | -0.001<br>(-0.024 - 0.021) | -0.155***<br>(-0.255 - -0.055) | -0.126***<br>(-0.216 - -0.037) |
| Observations | 6,555 | 6,517 | 5,130 | 6,422 | 6,517 |
| Outcome avg. | 0.229 | 0.359 | 0.068 | 0.246 | 0.180 |
| Est. p-value | 0.310 | 0.072 | 0.906 | 0.002 | 0.006 |

95% confidence intervals in parentheses below estimates. All columns estimated as Poisson regressions. Table reports average marginal effects. Standard errors clustered at block group-level. "Estimate p-value" in bottom row is exact p-value associated with the main coefficient estimate in column.
\*\*\* p<0.01, \*\* p<0.05, \* p<0.1

**Table 4.** Outcomes: Total arrests, drug arrests, and non-drug arrests per 100 in pop.

| VARIABLES | (1)<br>All | (2)<br>Drug | (3)<br>Not Drug |
| --- | --- | --- | --- |
| Post X 0-1 mi. X<br>MMC day | -0.045***<br>(-0.078 - -0.012) | -0.002<br>(-0.016 - 0.013) | -0.032**<br>(-0.061 - -0.002) |
| Post X 0-1 mi. X<br>All other days | -0.052***<br>(-0.082 - -0.023) | -0.015***<br>(-0.025 - -0.006) | -0.025***<br>(-0.043 - -0.006) |
| Observations | 46,189 | 42,807 | 46,037 |
| Outcome avg. | 0.150 | 0.038 | 0.115 |
| MMC day p-val. | 0.007 | 0.813 | 0.038 |
| All other days p-val. | 0.001 | 0.002 | 0.009 |

95% confidence intervals in parentheses below estimates. All columns estimated as Poisson regressions.
Table reports average marginal effects. Standard errors clustered at block group-level. "[estimate] p-value" in bottom row is exact p-value associated with the main coefficient estimates in column. These specifications are at block group-quarter-day of week level. All fixed effects are interacted with day-of-week indicator variables.
\*\*\* p<0.01, \*\* p<0.05, \* p<0.1

### Event studies

Event study results looked similar for each of our three main specifications (including total, drug, and non-drug arrest models) and none indicated a significant pre-trend. Each model saw an immediate but not statistically significant drop in arrest coefficients, then a steeper and significant drop during the second post period (6-12 months). Longer term impacts were less clear - each model saw arrest coefficients rise during the third post-period of more than 18 months, but this was only statistically significant for the total arrests category, and arrest rates still remained lower than in pre-periods in all categories (see Figure 2 for event study plots).

**FIGURE 2.**
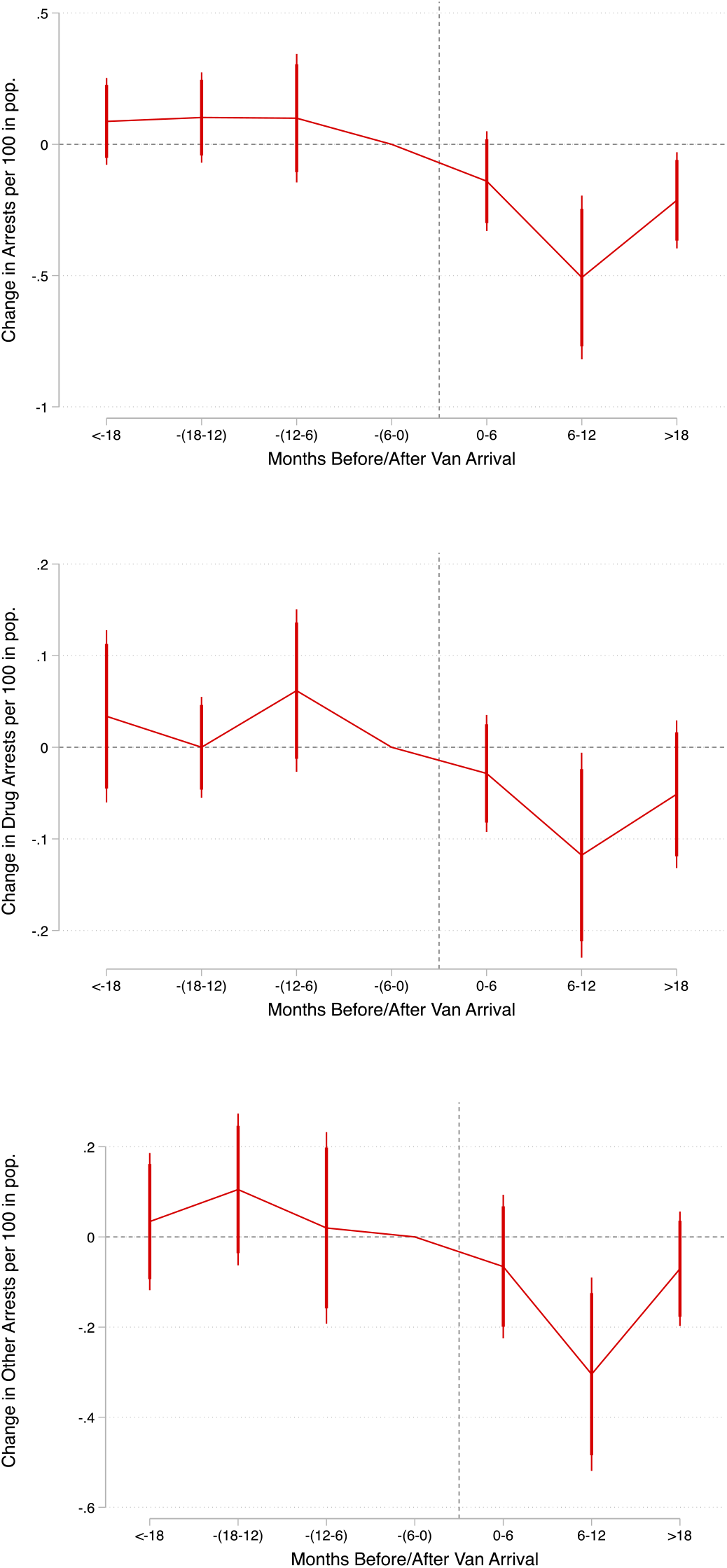
Event studies for total arrest, drug arrest, and non-drug arrest coefficients in intervention areas

### Robustness tests

Additional robustness models are reported in our supplementary materials (Appendices B and C). To briefly summarize, in testing for effects of MMCs at different distances, we found that results were generally similar when we coded block groups as treated if they were either within 0.5 or 1 mile from an MMC. When we extended beyond that range, to coding block groups 1.5 miles from the nearest MMC as “treated,” effect sizes were smaller and less significant (and one of 7 specifications suggests an increase in vehicular and public order arrests immediately outside treated areas – see Table B2 in supplementary materials). These results empirically support our choice of 1 mile as an appropriate “treatment” distance.

Other robustness tests aimed to construct a more comparable control group than simply using all block groups more than 1 mile from the nearest van. Our first attempt builds on the findings from the tests above. Namely, because there is a much more limited effect when we extend the treatment area to 1.5 miles, one might view the block groups that are 1-1.5 miles from the nearest MMC as a potential control group. The block groups may be more similar (simply by virtue of their propinquity) to the block groups within 1 mile but are also relatively unaffected by the vans. Therefore, in some tests, we restricted the sample to block groups within 1.5 miles and – as in our main analysis – coded block groups within 1 mile as treated units. The control group therefore exclusively consists of block groups 1-1.5 miles from the MMCs. Results are similar.

With a similar goal in mind, we also estimated propensity score weighted models and again observed similar results.

Finally, while Poisson models are sensible in light of the count data-nature of our outcome variables, we also estimated OLS models to ensure that results were not sensitive to model choice. Indeed, again, the basic conclusion of the OLS models were similar.

## DISCUSSION

### Summary

Our primary results suggest that MMCs yield lower overall arrest rates in their surrounding areas, with reductions primarily in drug, public order, vehicle, and violent crimes within a mile. The treatment areas, which had higher average drug arrest rates than the rest of the city prior to the arrival of MMCs, had lower average drug arrest rates than the rest of the city after the intervention.

MMCs saw sizable effects on most types of arrests in affected areas on days when they both were and were not active on site. These results suggest that MMCs decrease crime through their impact on individual drug use and drug related behaviors—not through general ecological or provider surveillance effects. One supplementary specification suggested that vehicular and public order charges increased slightly in the area between 1-1.5 miles from an MMC (see Supplementary Table B2). Given these crime types are highly prone to surveillance, these results could reflect a displacement of policing as much as a change in citizen behavior.

Results from supplementary robustness checks produced similar results to our main outcomes and indicated that effects were smaller and less significant as distance from the intervention increased. Together, these suggest that findings reflect more than analytic or measurement approach.

### Limitations

One limitation of the study is due to the arrest data’s treatment of drug charges, which do not identify arrests specific to opioids. Since buprenorphine only treats opioid use disorders, including other types of drug arrests introduces noise into the analysis, although this is more likely to have tempered than inflated effect estimates. A second limitation is the relatively short post-period. An extension of the analysis over time would shed light onto the longer-term implications of the intervention, which warrants attention since the most recent post-period in our event studies trended toward an increase in arrests (although these were still lower than pre-intervention arrest rates and largely non-significant).

## CONCLUSIONS

This study is the first of its kind to examine the impact of MMCs on local arrest rates. Outcomes indicate that there is no significant increase in crime of any kind in the area immediately surrounding the intervention, counter to NIMBY concerns that the introduction of harm reduction services could attract crime. In fact, MMCs may improve community safety, primarily by reducing criminalized behaviors related to addiction (i.e. illicit drug use and possession), but also by impacting peripheral offenses including public order, vehicular, and violent crime.

Mobile buprenorphine provision expands access to a gold standard treatment whose potential has long been hampered by social and bureaucratic obstacles that hinder access for those who need it most (16–18). MMC expansion could promote both health equity and community safety.

## Data Availability

All data used in the present study are available online at: City of Pittsburgh; 2023; Pittsburgh police arrest data; Western Pennsylvania Regional Data Center; doi: d809c36f-28fe-40e6-a33e-796f15c66a69.

https://catalog.data.gov/dataset/pittsburgh-police-arrest-data

