## Supporting Information for "There Goes the Neighborhood? The Public Safety Enhancing Effects of a Mobile Harm Reduction Intervention"

**Appendix A: Intervention Details**

The assessed intervention provides low-barrier buprenorphine access from mobile health clinics attending established Syringe Service Program sites. The goal of the service is to reduce the limitations on the availability of buprenorphine in Allegheny County, given the unprecedented rate of morbidity and mortality among people who use opioids. By eliminating cost and location barriers from the operation of buprenorphine services, Prevention Point Pittsburgh (PPP) worked to create a patient-centric and community-based alternative to existing MOUD access structures. Given the current context, in which the drug supply is increasingly potent and unpredictable, low-barrier access to MOUD was deployed to extend the reach of a proven intervention.

The non-punitive nature of service delivery is reflective of the intervention’s theoretical foundation. Appointments are not required, and patients are always welcome to return to treatment after an absence regardless of their reason. Urinalysis drug screening is primarily used to check for buprenorphine adherence and any unexpected results are handled in discussions rather than addressed by automatic consequences such as termination of treatment.

The team of providers for the intervention include a social worker, and a team of contracted MOUD providers from a regional health system. On a typical day the low barrier buprenorphine service is operated by the Social Worker and one or two contracted providers based on expected volume. The contracted provider team includes physicians, physicians assistants, and registered nurse practitioners.

Upon initiation, clients meet with the social worker to register, prepare for the visit, and identify their treatment needs and goals. They then meet with the medical provider to discuss substance use history and eligibility for starting and/or continuing buprenorphine services. If appropriate, the provider submits a prescription to the patient’s pharmacy of choice for same-day pickup. If the patient is receiving assistance with the cost of pharmacy fills, partner pharmacies in the vicinity of each site are able to bill the program directly. Telehealth visits are conducted as needed, but in-person visits are highly encouraged given the other resources available at the PPP sites.

Services are delivered from a large cargo van which has been converted to include the core features of a clinic room. The van features an exam table, restroom with non-flush toilet, sink, refrigerator, storage, temperature control, accessory power via a generator, supply storage, a small countertop, alternate lighting options, and a step-in side access door. The vehicle does not require a commercial driver’s license to operate, as it is not considered oversized and can fit in a standard parking space. A WiFi hotspot allows for the use of an electronic health record.

The service operates Tuesdays - Thursdays at PPP’s van-based syringe service program locations in the City of Pittsburgh. Sites are open for 3 hours each week.

**Appendix B:** Primary outcomes at other distances from MMCs

| **Table B1.** Outcomes: Total arrests and drug arrests per 100 in pop. | | |
| --- | --- | --- |
|  | (1) | (2) |
| VARIABLES | All | Drug |
| Post*0-0.5 Mi. | -0.351*** | -0.184*** |
|  | (0.104) | (0.052) |
| Post*0.5-1 Mi. | -0.243*** | -0.162** |
|  | (0.090) | (0.075) |
| Post*1-1.5 Mi. | -0.035 | 0.003 |
|  | (0.088) | (0.076) |
| Observations | 6,630 | 6,417 |
| Outcome avg. | 0.975 | 0.436 |
| Omitted category: block groups >1.5 mi. from vans. All columns estimated as Poisson regressions. Table reports average marginal effects. Standard errors in parentheses. | | |

**Table B2.** Outcomes: Other Arrest Categories per 100 in pop.

|  | (1) | (2) | (3) | (4) | (5) |
| --- | --- | --- | --- | --- | --- |
| VARIABLES | Property | Violent | Weapon | Pub. Order | Vehicle |
| Post*0-0.5 Mi. | 0.009 | -0.083 | -0.123** | -0.211*** | -0.132*** |
|  | (0.060) | (0.139) | (0.057) | (0.060) | (0.049) |
| Post*0.5-1 Mi. | -0.019 | -0.044 | -0.064 | -0.112** | -0.034 |
|  | (0.050) | (0.137) | (0.056) | (0.050) | (0.045) |
| Post*1-1.5 Mi. | 0.026 | 0.254 | -0.056 | 0.153* | 0.255*** |
|  | (0.074) | (0.171) | (0.057) | (0.081) | (0.098) |
| Observations | 6,607 | 6,554 | 5,167 | 6,452 | 6,547 |
| Outcome avg. | 0.415 | 0.615 | 0.147 | 0.447 | 0.370 |
| Omitted category: block groups >1.5 mi. from vans. All columns estimated as Poisson regressions. Table reports average marginal effects. Standard errors in parentheses. | | | | | |

**Appendix C:** Robustness tables for total, drug, and all other arrest analyses with different spatial specifications.

| **Table C1**. Outcomes: Total arrests per 100 in pop | | | | | | | |
| --- | --- | --- | --- | --- | --- | --- | --- |
|  | (1) | (2) | (3) | (4) | (5) | (6) | (7) |
|  | Treatment= | Treatment= | Treatment= | Sample= | Prop. Score* |  | OLS |
| VARIABLES | 0-0.5 mi | 0-1 mi | 0-1.5 mi | 0-1.5 mi | IPW | OLS | +IPW |
| Post X 0-0.5 mi. | -0.377*** |  |  |  |  |  |  |
|  | (-0.621 - -0.133) |  |  |  |  |  |  |
| Post X 0-1 mi. |  | -0.358*** |  | -0.699*** | -0.137*** | -0.436*** | -0.216*** |
|  |  | (-0.557 - -0.158) |  | (-1.072 - -0.326) | (-0.237 - -0.038) | (-0.651 - -0.221) | (-0.355 - -0.077) |
| Post X 0-1.5 mi. |  |  | -0.085 |  |  |  |  |
|  |  |  | (-0.289 - 0.119) |  |  |  |  |
| Observations | 6,612 | 6,612 | 6,612 | 2,831 | 6,536 | 6,612 | 6,536 |
| Outcome avg. | 1.049 | 1.049 | 1.049 | 1.731 | 0.734 | 1.049 | 0.734 |
| Est. p-value | 0.002 | 0.000 | 0.413 | 0.000 | 0.007 | 0.000 | 0.002 |

Omitted category: block groups >1.5 mi. from vans. All columns estimated as Poisson regressions. Table reports average marginal effects. Standard errors in parentheses.

**Table C2**. Outcomes: Total drug arrests per 100 in pop.

|  | (1) | (2) | (3) | (4) | (5) | (6) | (7) |
| --- | --- | --- | --- | --- | --- | --- | --- |
|  | Treatment= | Treatment= | Treatment= | Sample= | Prop. Score* |  | OLS |
| VARIABLES | 0-0.5 mi | 0-1 mi | 0-1.5 mi | 0-1.5 mi | IPW | OLS | +IPW |
| Post X 0-0.5 mi. | -0.072** |  |  |  |  |  |  |
|  | (-0.133 - -0.011) |  |  |  |  |  |  |
| Post X 0-1 mi. |  | -0.087*** |  | -0.202*** | -0.047** | -0.259*** | -0.119** |
|  |  | (-0.151 - -0.023) |  | (-0.331 - -0.073) | (-0.087 - -0.007) | (-0.398 - -0.120) | (-0.212 - -0.026) |
| Post X 0-1.5 mi. |  |  | -0.033 |  |  |  |  |
|  |  |  | (-0.091 - 0.025) |  |  |  |  |
| Observations | 6,384 | 6,384 | 6,384 | 2,774 | 6,308 | 6,612 | 6,536 |
| Outcome avg. | 0.257 | 0.257 | 0.257 | 0.417 | 0.202 | 0.248 | 0.195 |
| Est. p-value | 0.021 | 0.008 | 0.270 | 0.002 | 0.022 | 0.000 | 0.012 |

Omitted category: block groups >1.5 mi. from vans. All columns estimated as Poisson regressions. Table reports average marginal effects. Standard errors in parentheses.

**Table C3**. Outcomes: Total non-drug arrests per 100 in pop.

|  | (1) | (2) | (3) | (4) | (5) | (6) | (7) |
| --- | --- | --- | --- | --- | --- | --- | --- |
|  | Treatment= | Treatment= | Treatment= | Sample= | Prop. Score* |  | OLS |
| VARIABLES | 0-0.5 mi | 0-1 mi | 0-1.5 mi | 0-1.5 mi | IPW | OLS | +IPW |
| Post X 0-0.5 mi. | -0.197** |  |  |  |  |  |  |
|  | (-0.352 - -0.041) |  |  |  |  |  |  |
| Post X 0-1 mi. |  | -0.179*** |  | -0.335*** | -0.070* | -0.177*** | -0.097** |
|  |  | (-0.302 - -0.057) |  | (-0.551 - -0.118) | (-0.150 - 0.011) | (-0.287 - -0.067) | (-0.173 - -0.020) |
| Post X 0-1.5 mi. |  |  | -0.051 |  |  |  |  |
|  |  |  | (-0.211 - 0.109) |  |  |  |  |
| Observations | 6,593 | 6,593 | 6,593 | 2,831 | 6,517 | 6,612 | 6,536 |
| Outcome avg. | 0.803 | 0.803 | 0.803 | 1.323 | 0.540 | 0.801 | 0.538 |
| Est. p-value | 0.013 | 0.004 | 0.531 | 0.003 | 0.089 | 0.002 | 0.013 |

Omitted category: block groups >1.5 mi. from vans. All columns estimated as Poisson regressions. Table reports average marginal effects. Standard errors in parentheses

***Additional information on propensity score matching**

We conducted k-nearest-neighbors propensity score matching based on block group demographics, identifying 10 matches for each of the treatment units to use as a control group. Factors used in matching include age, total population, ratio of income to poverty level, per capita income in the last 12 months, occupancy status and residency tenure. Any control observations outside a 40-90% range of treatment likelihood based on these factors (compared to the actual treatment areas) were dropped.
